# Trend and Weightage of Problem-based Questions in Undergraduate Pharmacology Written Question Papers of Bangladesh

**DOI:** 10.1101/2022.02.02.22270345

**Authors:** Fatema Johora, Asma Akter Abbasy, Sabiha Mahboob, Fatiha Tasmin Jeenia, Jannatul Ferdoush, Md. Sayedur Rahman

## Abstract

**Background:** The key objective of pharmacology education is to make graduates competent enough to prescribe safely and effectively. There is worldwide speculation of inadequacy of pharmacology education to achieve the expected learning outcomes. Problem-based pharmacotherapy is considered as one of the crucial way to prepare future physicians as rational prescribers. As assessment drives learning priorities, adequate weightage on problem-based questions in undergraduate pharmacology examination might be helpful.

**Materials and Methods:** This descriptive cross-sectional study was conducted to analyze the undergraduate pharmacology written question papers (SAQ) of MBBS curriculum of 07 different universities (Bangladesh University of Professionals, University of Dhaka, University of Chittagong, University of Rajshahi, Shahjalal University of Science and Technology, University of Science and Technology, Chittagong and Gono Bishwabidyalay) of Bangladesh in last 10 years (January 2010 to November 2019). Total 131 question papers were collected, and trend and weightage of problem-based questions were evaluated.

**Results:** Problem-based questions have been reduced dramatically over the decade and Mean percentage of marks allocated for problem-based questions was 1.2±1.3 over last years. There was significant difference (<0.00001) of weightage of problem-based questions among different universities of Bangladesh. Highest presence of problem-based question was observed in Gono Bishwabidyalay (GB), followed by Shahjalal University of Science and Technology (SUST), University of Rajshahi (RU) and Bangladesh University of Professionals (BUP) but there was not a single problem-based question in University of Dhaka (DU), University of Chittagong (CU) and University of Science and Technology (USTC) over 10 years period.

**Conclusion:** Current study revealed negligible presence of problem-based questions in undergraduate pharmacology written question papers of Bangladesh over 10 years period.

## INTRODUCTION

Assessment is a fundamental component of medical education as it drives students learning and certification.^1^ It can be formative or summative. Formative assessment reinforces students’ intrinsic motivation to learn and inspire them to set higher standards for themselves.^2, 3^ On the other hand, summative assessment is intended to provide professional self regulation and accountability which may also act as a barrier to further practice or training. In summative assessment, students tend to study that which they expect to be tested on and it influence learning even in the absence of feedback.^4, 5^ Written assessment has been used for almost as long as medicine has been taught. It has historically been the cornerstone of testing doctors’ and medical students’ knowledge. In modern times, it is still used throughout the world, at all stages of medical training, principally as a means of testing knowledge.^6^ Various types of questions can be used like MCQ, SAQ, MEQ and SEQ.^7^ In the past, long essay questions were commonly used in written examination to assess the cognitive ability of students. But this traditional essay question could not yield the expected answers and wide variation in student’s interpretation was noted. Therefore it is showed limited validity, poor reliability and less objectivity.^8, 9^ On the other hand, SAQ direct the students towards a precise and specific response and provide greater objectivity, reliability and their range of content area testes is extended. There are several types of SAQ questions including completion items, definitions, unique answer type, label/draw diagram, numerical problems, open ‘SAQs’ and problem-based questions. Problem-based questions is useful to assess whereby medical students can apply and / or transform their knowledge and understanding in order to diagnose and treat clinical conditions which they are supposed to perform in their practice in upcoming days.^7, 10^

The goal of medical education is to prepare graduates who are competent enough to prescribe safely and effectively.^11^ A thorough understanding of pharmacology is needed for effective pharmacotherpy and prescribing of medications.^12, 13^ It is expected that pharmacology teaching program should be able to develop a foundation to enable them to appraise new medicines throughout professional career. In many medical schools, pharmacotherapy teaching is characterized by the transfer of knowledge about drugs, rather than by skill to treat patients. Actually pharmacotherapy is a skill; it is more than knowledge alone. Although World Health Organization (WHO) urges for outcome-based learning in pharmacology, focusing on practice rather than on theory^14^, studies show growing concerns about inadequacy of pharmacology education to prepare future physicians as rational prescribers.^15, 16, 17^ Problem-based pharmacotherapy teaching is the best way to make rational prescribers. Problem-based curriculum is not necessary for teaching pharmacotherapy. It is possible to integrate problem-based pharmacotherapy in a traditional curriculum. Introduction of few hours of problem-based learning within the limits of current curriculum, problem-solving exercises and problem-solving questions in different parts of standard assessment systems would be helpful to achieve this goal.^14^

Bangladesh inherited medical education program from the British, and then Pakistan.^18^ In 1988, first documented and reformed curriculum for undergraduate medical program was introduced, then subsequently revised in 2002 and 2002.^19, 20^ Several studies were conducted in Bangladesh to evaluate pharmacology education (curriculum, textbooks and question papers) from different perspectives.^21, 22, 23, 24, 25, 26^ In this backdrop, current study was conducted to evaluate pharmacology written question papers of last 10 years with the attempt to find out the weightage and trend of problem-solving questions in the written assessment system.

## MATERIALS & METHODS

The objective of this study was to assess the weightage and trend of problem-based questions in undergraduate pharmacology written question papers of Bangladesh.

### Study Design and Procedure

A descriptive cross-sectional study was designed to meet the study objective and was conducted from January 2021 to March 2021. Ethical approval was taken from the Institutional Review Board (IRB) of Combined Military Hospital (CMH) Bogura, Bangladesh. Pharmacology written question papers (SAQ) of last 10 years (January 2010 to November 2019) of all 7 universities offering MBBS degree (Bangladesh University of Professionals, University of Dhaka, University of Chittagong, University of Rajshahi, Shahjalal University of Science and Technology, University of Science and Technology, Chittagong and Gono Bishwabidyalay) were collected and included in the study for analysis. Total 131 question papers were collected for review. Researchers thoroughly screened the question papers, sorted out problem-solving questions and calculated the weightage. The Pharmacology written question papers (SAQ) contain total 84 marks with options, where students need to answer a maximum of 70 marks. Weightage was calculated as the number reflecting problem-solving question out of 84 marks.

### Statistical analysis

Data was compiled, presented and and analyzed using Microsoft Excel 2007, and was expressed as mean percentage (standard deviation). One Way Analysis of Variance (ANOVA) was done to determine the significance of difference between the mean percentages. Statistical analysis was performed at a 95% confidence interval and significance was determined at p< 0.05.

## RESULTS

Total 131 SAQ papers of undergraduate pharmacology written question papers dated from January 2010 to November 2019 were analyzed. There was not a single problem-based question in University of Dhaka (DU), University of Chittagong (CU) and University of Science and Technology (USTC) over 10 years period. Mean percentage of marks allocated for problem-based questions was 1.2±1.3 over last years. Highest presence of problem-based question was observed in Gono Bishwabidyalay (GB) **3.9±3.7**, followed by Shahjalal University of Science and Technology (SUST), University of Rajshahi (RU) and Bangladesh University of Professionals (BUP). And statistically significant difference (<0.00001) was observed among seven universities (Table I).

**Table I:** Weightage of Problem based questions.

| DU<br>(Mean ±<br>SD) | CU<br>(Mean ±<br>SD) | RU<br>(Mean ±<br>SD) | SUST<br>(Mean ±<br>SD) | USTC<br>(Mean ±<br>SD) | GB<br>(Mean ±<br>SD) | BUP<br>(Mean ±<br>SD) | Total<br>(Mean ±<br>SD) | P value |
| --- | --- | --- | --- | --- | --- | --- | --- | --- |
| 0.00± 0.00 | 0.00±0.00 | 1.7±3.2 | 2.6±4.2 | 0.00±0.00 | 3.9±3.7 | 0.3±1.0 | 1.2±1.3 | $<0.00001$ |

Data expressed as mean percentage (SD) and analyzed by One Way Analysis of Variance (ANOVA), $p < 0.05$ -Significant

**Figure 1** represented the trend of problem-based questions (i.e. mean percentage) in pharmacology written question papers throughout the last ten years. The percentage of problem-based question had been reduced dramatically over the years almost climbing down to nil.

**Figure 1:**
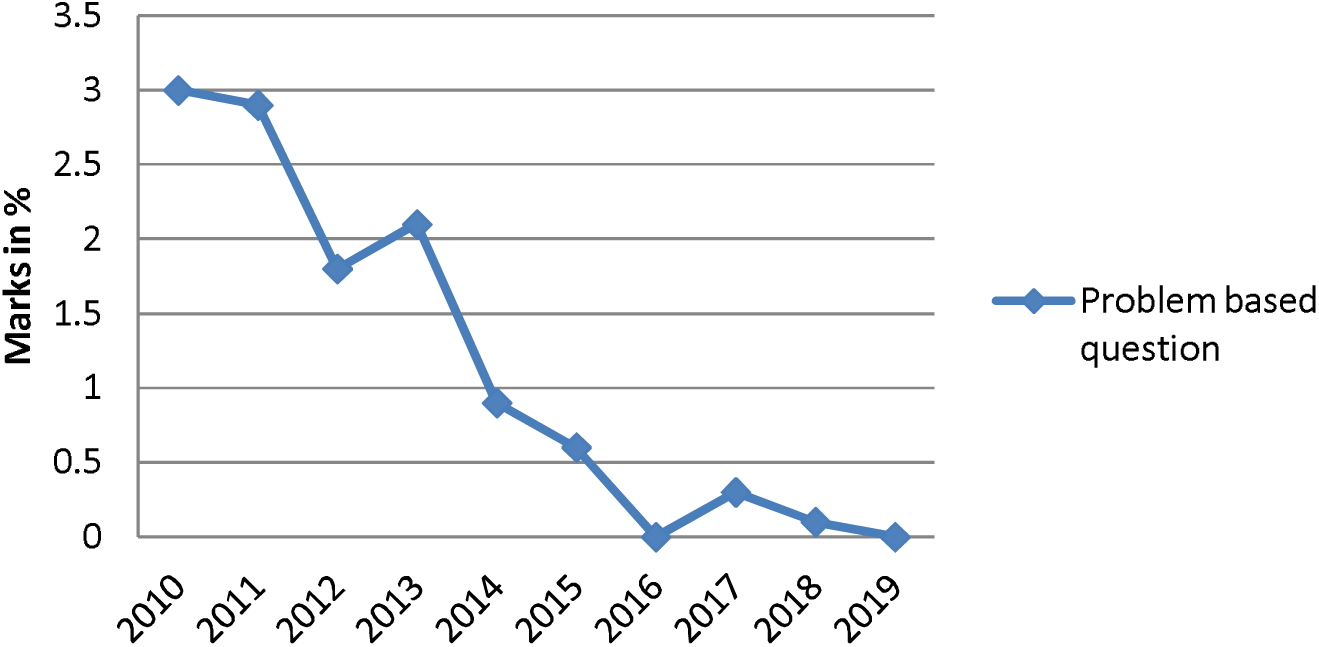
Trend of problem based question in pharmacology question papers.

## DISCUSSION

Prescribing is a complex process involving a mixture of knowledge, attitude and skills. Preparing graduates to be rational prescribers is one of the greatest challenges of undergraduate pharmacology education.^27^ It has been generally felt that pharmacology course in undergraduate medical education is not adequate to keep pace with the rapid changes and requirements of clinical practice. Traditionally, it has focused more on factual information with little or no emphasis on practical aspects of prescribing. Effectiveness of current pharmacology eduation as a foundation to make graduate competent enough to prescribe safely and effectivly is rather questionable.^28, 29^ Problem-based pharmacotherapy has been considered as the first step in a long process of changing from traditional teaching towards an integrated curriculum with problem-based learning with the aim to make graduates as competent prescribers.^14^ Current study was conducted in this context to assess the weightage and trend of problem-based questions in undergraduate pharmacology written question papers of different universities of Bangladesh for last 10 years.

The term ‘medical problem-solving’ is most often used to refer to that process by which physicians arrive at a diagnosis, select a management strategy, initiate remedial action, monitor its effectiveness and modify their interventions accordingly. The situations in which this process may be used range from individual patient discomforts to community or national health problems. Assessment of problem-solving skills is best done by placing an individual in a problematical situation and observing his performance.^30^ Problem based questions are for students to assess their knowledge and understanding level through which they can solve problem in a novel situation.^31^ Current research found that percentage of problem based question had reduced dramatically over the last 10 years and this finding was concurrent to previous two studies conducted in Bangladesh^24, 26^ although there is already identified need by pharmacology educators of the country to incorporate problem based questions in undergraduate written exams.^32^ Regarding weightage of problem based questions, another point is significant difference (<0.00001) was observed among different universities in current study. Furthermore, a study has revealed the effectiveness of problem-based learning in context of Bangladesh.^33^ A blend of problem based learning along with conventional classes in pharmacology classroom can trigger the student’s efficiency and prescribing behavior in a prudent way. Therefore, more highlight on problem based teaching learning and eventually on question set up in pharmacology is recommended.

Assessment is the central feature of medical education. Changing the examination system without changing the curriculum had a much more profound impact upon the nature of learning than changing the curriculum without altering the examination.^6, 7^ Incorporation of problem-based questions in undergraduate pharmacology examination will motivate students about problem-based pharmacotherapy as examination is the driving force for learning in medical education.^34^

## CONCLUSION

Current study showed negligible presence of problem based questions in undergraduate pharmacology written question papers in last 10 years and inter-university variation of weightage was observed. Undergraduate medical students who have been taught by same curriculum could have different learning outcomes as assessment determines learning priority. A clear direction about weightage of problem based questions is needed to spell out in the undergraduate pharmacology curriculum. A test blueprint and question bank might be helpful as a reference tool for the question setters and moderators to prepare question for achieving expected learning outcomes.

## Data Availability

All data produced in the present study are available upon reasonable request to the authors

## REFERENCE

1. Norcini JJ. Peer assessment of competence. Med Educ 2003; 37:539–43. http://dx.doi.org/10.1046/j.1365-2923.2003.01536.x

2. Ferris H, Flynn DO. Assessment in Medical Education; What Are We Trying to Achieve? International Journal of Higher Education, 2015; 4 (2): 139–144.

3. Leung WC. Competence based medical training: Review. BMJ 2002; 325:693–6. http://dx.doi.org/10.1136/bmj.325.7366.693

4. Norcini JJ, McKinley DW. Assessment methods in medical education. Teaching and Teacher Education, 2007; 23: 139–150

5. Lockyer J, Carraccio C, Chan MK, Hart D, Smee S, Touchie C and et al. Core principles of assessment in competency-based medical education, Medical Teacher, 2017; 39:6, 609-616

6. Hayes K. Written assessment. In: Walsh K, ed. Oxford Textbook of Medical Education. Oxford University Press, 2013.

7. Sood R. Assessment in medical education: Trends and tools. All India Institute of Medical Science, New Delhi, India, 1995.

8. Cunningham GK. Assessment in the classroom: Constructing and interpreting tests. Falmer Press, London, 1998.

9. Feletti GI, Smith EK. Modified Essay Questions: Are they worth the effort? Medical Education, 1986; 20 (2):126–132.

10. Ellingon H. Short answer questions. In: Teaching and learning in higher education. Scottish Central Institutions Committee Educational development. Aberdeen, UK, 1987.

11. World Medical Association (WMA). WMA Statement on Medical Education. 2020. Available at: https://www.wma.net/policies-post/wma-statement-on-medical-education/ [Accessed on 02/11/2020]

12. Engels F. Pharmacology education: Reflection and challenges. European Journal of Pharmacology. 2018; 833: 392–95.

13. Maxwell, S., Walley, T., 2003. Teaching safe and effective prescribing in UK medical schools: a core curriculum for tomorrow’s doctor. British Journal of Clinical Pharmacology, 55, pp. 496–503.

14. World Health Organization (WHO). Teacher’s Guide to Good Prescribing. World Health Organization, Geneva, Switzerland. 2001. Available at: http://apps.who.int/iris/bitstream/10665/67010/1/WHO_EDM_PAR_2001.2.pdf [Accessed on 02/11/2021]

15. Tobaiqy M, McLay J, Ross S. Foundation year 1 doctors and clinical pharmacology and therapeutics teaching. A retrospective view in light of experience. Br J ClinPharmacol. 2007, 2007: 363–372.

16. Heaton A, Webb DJ, Maxwell SRJ. Undergraduate preparation for prescribing: the views of 2413 UK medical students and recent graduates. Br J ClinPharmacol. 2008, 2008: 128–134.

17. loyd LH, Hinton T, Bullock S, Babey A, Davis E, Fernandes L, et al. An evaluation of pharmacology curricula in Australian science and health-related degree programs. BMC Medical Educaton. 2013; 13.

18. Faiz MA. Medical education in Bangladesh-Is there room for improvement? JCMCTA. 2007; 18: 1–3.

19. Bangladesh Medical & Dental Council (BMDC). Curriculum for Undergraduate Medical Education in Bangladesh – updated 2002. Bangladesh Medical & Dental Council (BMDC), 2002, Dhaka, Bangladesh. Available at: http://bmdc.org.bd/mbbs-curriculum-update-2002/ [Accessed on 02/11/2021]

20. Bangladesh Medical & Dental Council (BMDC). Curriculum for Undergraduate Medical Education in Bangladesh – updated 2012. Bangladesh Medical & Dental Council (BMDC), 2012, Dhaka, Bangladesh. Available at: http://bmdc.org.bd/mbbs-curriculum-update-2012/ [Accessedon 02/11/2021]

21. Begum M, Rahman MS, Islam AFMS, Khan IA, Akhter N. Eleven Years of the Undergraduate Medical Curriculum 1988: review on the changes in Pharmacology written questions. Bangladesh J PhysiolPharmacol. 1999; 15: 27–30.

22. Karim A, Haque M. 1996. Assessment system in Pharmacology-Does it Reflect Educational objectives and Community Health Needs? Bangladesh J Physiol Pharmacol. 1996; 12: 65–67.

23. MS Rahman, S Huda. Antimicrobial resistance and related issues: An overview of Bangladesh situation. Bangladesh Journal of Pharmacology. 2014;9, 218–224.

24. Chowdhury DK, Saha D, Talukder MH, Habib MA, Islam A, Ahmad MR, Hossin MI. Evaluation of Pharmacology Written Question Papers of MBBS Professional Examinations. Bangladesh Journal of Medical Education. 2017; 8: 12–17.

25. Johora F, Rahman MS. Pharmacology education in the perspectve of pharmaceutical promotion: Bangladesh experience. Bangabandhu Sheikh Mujib Med Univ J. 2019; 12: 128–132.

26. Nahar N, Kutubi A, Khan TH, Banu LA, Jahan I, Badhon NM. Analysis of undergraduate pharmacology written question of different universities I Bangladesh. J Shaheed Suhrawardy Med Coll 2017: 9 (1) 13–17.

27. Ross S, Maxwell S. Prescribing and the core curriculum for tomorrow’s doctors: BPS curriculum in clinical pharmacology and prescribing for medical students. Br J Clin Pharmacol. 2012; 74(4):644–661.

28. Brinkman DJ, Tichelaar J, Graaf S, Otten RHJ, Richir MC, van Agtmael MA. Do final-year medical students have sufficient prescribing competencies? A systematic literature review. Br J Clin Pharmacol. 2018; 84(4):615–635.

29. Heaton A, Webb DJ, Maxwell SRJ. Undergraduate preparation for prescribing: the views of 2413 UK medical students and recent graduates. Br J Clin Pharmacol 2008; 66:128–134

30. McGuire CH. Assessment of Problem-Solving Skills, 1. Medical teacher, 1980; 2 (2): 74–79.

31. Tabish SA. Assessment methods in medical education. Int J Health Sci. 2008; 2:3–7.

32. Chowdhury DK, Saha D, Habib MA. Teachers’ Opinion about Pharmacology Written Question Papers of MBBS Professional Examinations. Bangladesh Journal of Medical Education 2021, 12(1), 40–49.

33. Jeenia FT, Hoque A, Khanom M, Jahangir SM, Hoque R, Parveen K, Ferdoush J, Ata M, Tanin MJU. Introducing Problem-Based Learning as an Effective Learning Tool to Medical Students: An Approach in Bangladesh. Bangladesh Journal of Medical Education 2021; 12(1): 22–31.

34. Al-Kadri HM, Al-Monary MS, Roberts C, Van Der Vluuten CPM. Exploring assessment factors contributing to students’ study strategies: Literature review. Med Teach.2012; 34: 42–50.

